# No Evidence for Reduced Hospital Admissions or Increased Deaths from Stroke or Heart Attack During COVID-19

**DOI:** 10.1101/2020.06.08.20119636

**Authors:** Katharine Reeves, Samuel Watson, Tanya Pankhurst, Kamlesh Khunti, Suzy Gallier, Magdalena Skrybant, Peter J Chilton, Richard J Lilford

## Abstract

Articles in the UK press have claimed that hospital admissions for heart attack and stroke have declined during the COVID-19 pandemic. However, data from the West Midlands Ambulance Service have not shown any reduction in call-outs for patients with stroke or ST-Elevation Myocardial Infarction. This study examined data from University Hospital Birmingham NHS Foundation Trust, comparing admissions from week 1 of 2016 to week 17 of 2019, with the same period in 2020, pre- and post-lockdown. The results showed that there was no evidence of a reduction in the overall mean number of admissions of patients with these conditions in the post-lockdown period.

---

Claims that hospital admissions for heart attack and stroke have declined over the COVID-19 pandemic have been made in at least 16 newspaper articles in the UK (Table 1). In contrast, data from the West Midlands Ambulance Service (WMAS) have not reported any reduction in call-outs for patients with stroke or ST-Elevation Myocardial Infarction (STEMI).^1^ However, not all admissions with these diseases arrive via the Accident and Emergency (A&E) department. Furthermore, a study from Italy showed around a 30% reduction in acute coronary syndrome related hospital admission during the COVID-19 crisis.^2^ We therefore examined admissions to University Hospitals Birmingham NHS Foundation Trust, which is the UK’s largest hospital group and lies within the WMAS footprint.

**Table 1.**
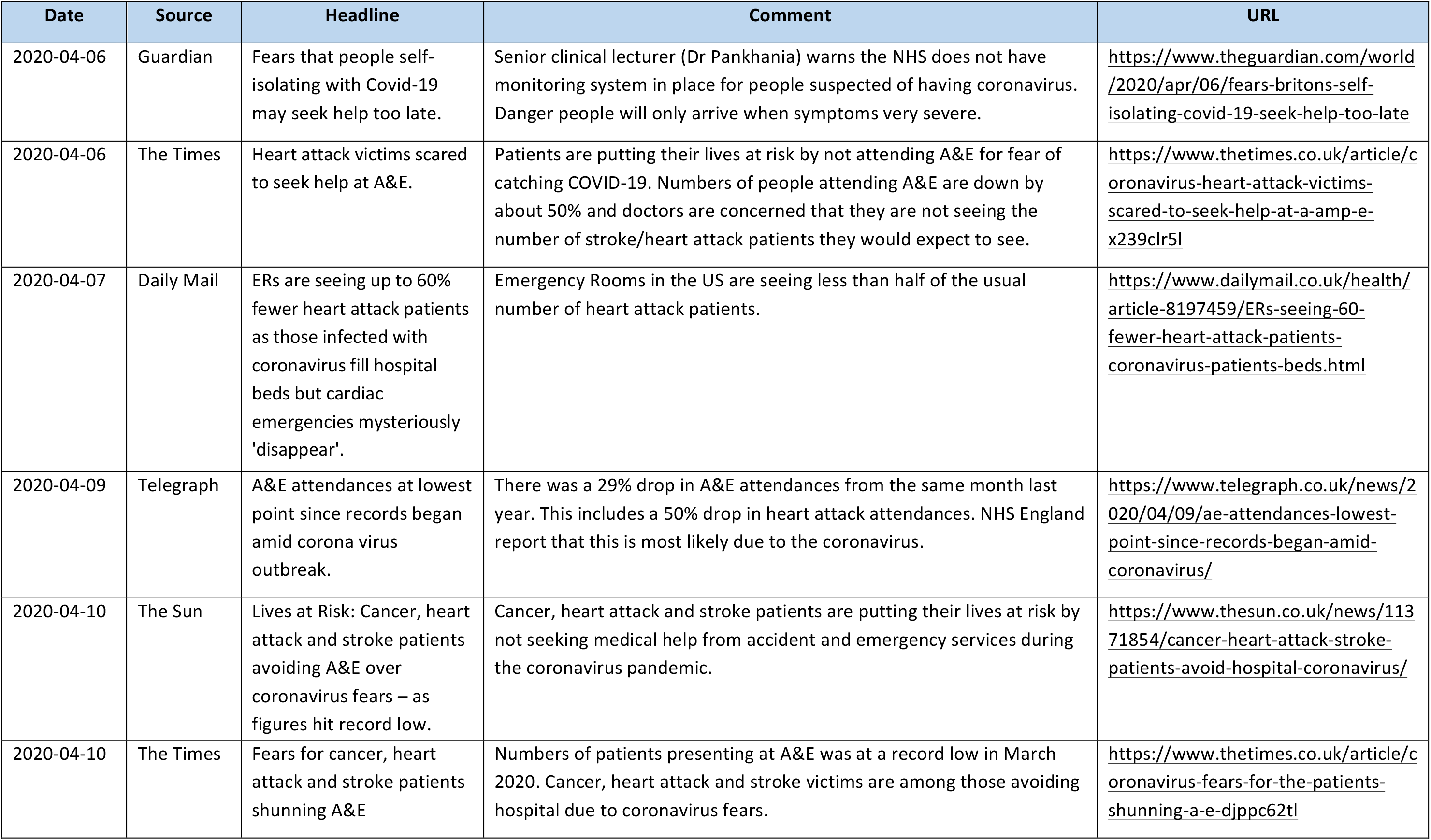
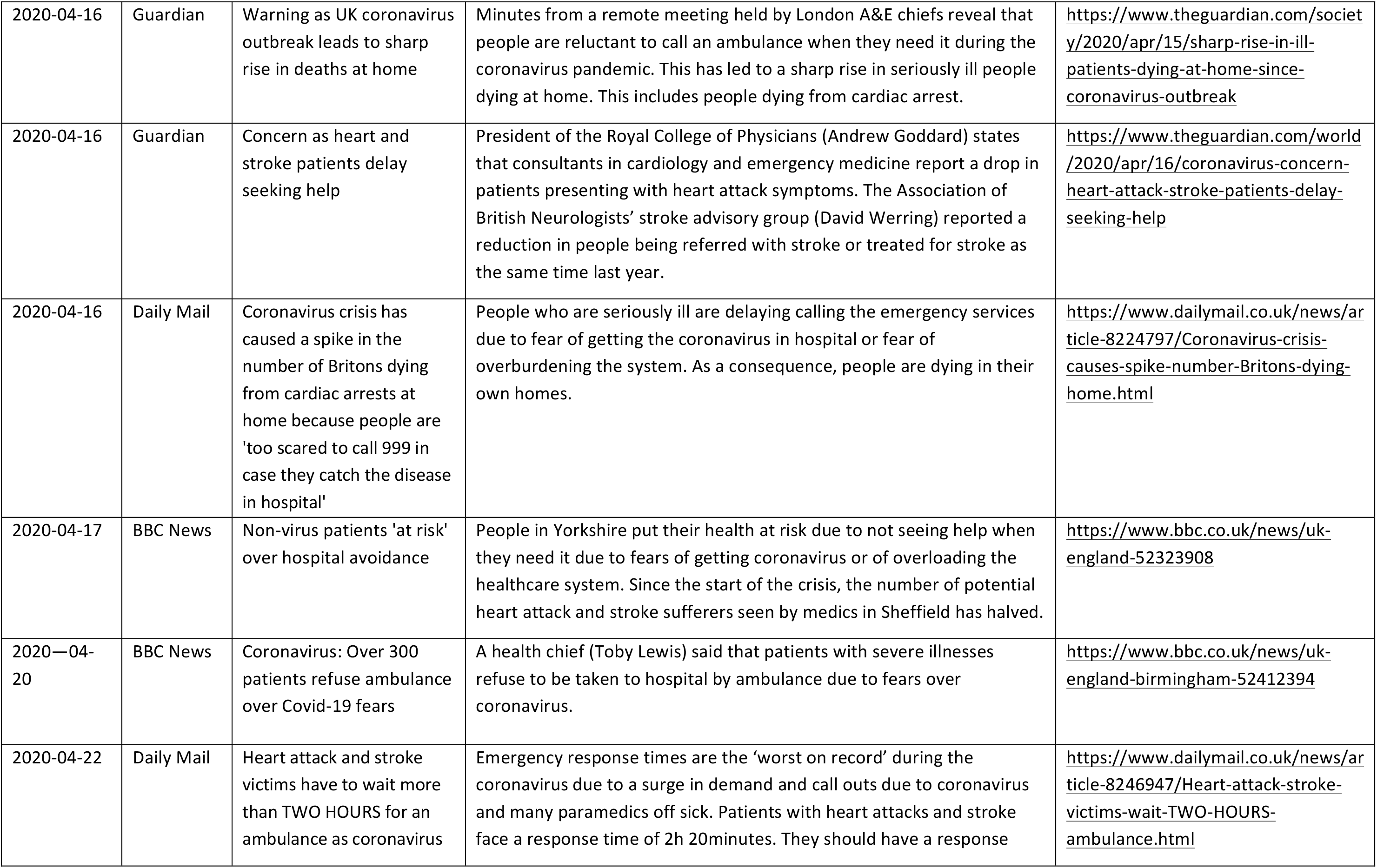
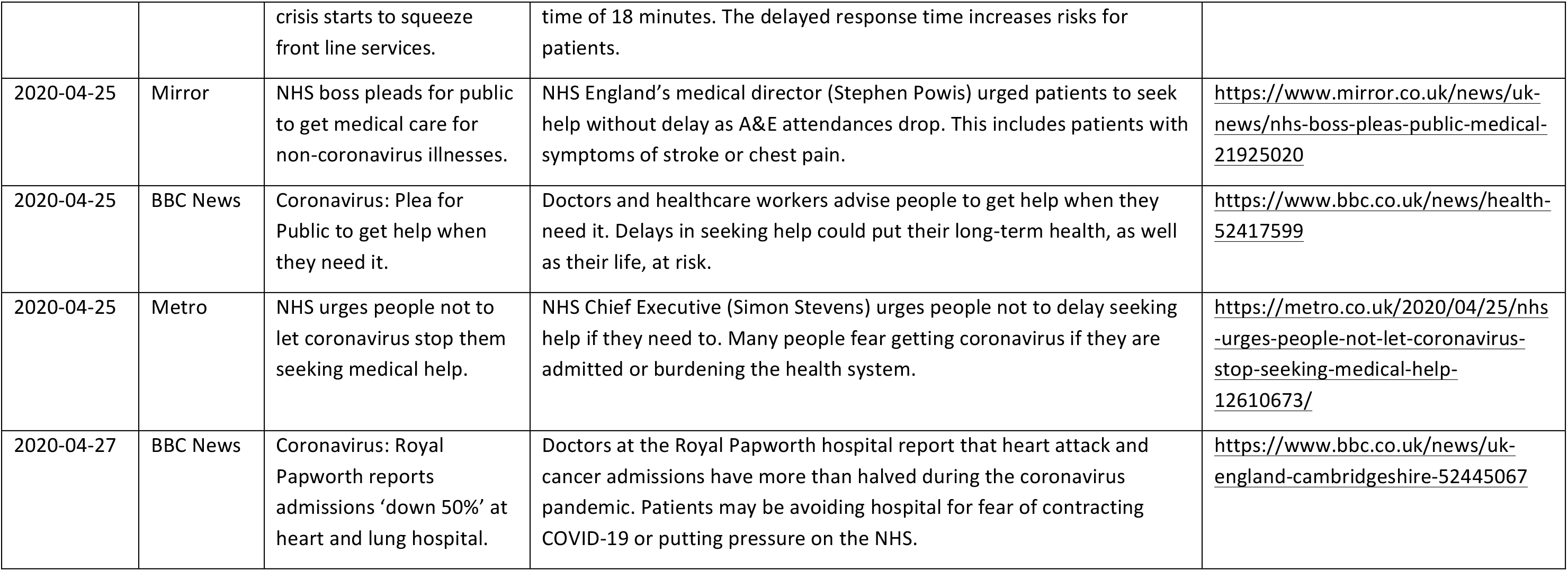
Newspaper reports on declining access to hospital care for stroke and heart attack

We examined cases admitted with STEMI (Codes I21·0 to I21·3 and I21·9) and stroke (Codes I61·0 to I61·9 and I63·0 to 163·9) from week 1 of 2016 to week 17 of 2019 and compared to the same period in 2020 pre and post lockdown. We examined three end-points: total admissions, admissions from A&E, and seven-day in-hospital mortality. The figure shows the number of admissions over time and proportionate differences from the 2016-2019 average. Small reductions in weekly admissions were observed for STEMI (−4·2, 95% CI [-10·6, 2·1], p=0·17) and stroke (−4·4 [-10·8, 2·0], p=0·15), but there was little evidence that this represents a shift in the overall mean in the post-lockdown period. We found little evidence of a difference in the number of patients being admitted through the emergency department (STEMI: -3·6 percentage points [pp], 95% CI [2·1, -9·3], p=0·19; stroke: -5·0 pp, 95% CI [1·7, -11·7], p=0·19) or seven-day in-hospital mortality (STEMI: -0·0 pp, 95% CI [1·4, -1·4], p=1·00; stroke: -1·6 pp, 95% CI [1·1, -4·3], p=0·21). Examination of the long run series in the Figure shows that there were larger spikes and troughs over the entire observation period, than those observed after lockdown occurred.

In summary, despite reported media claims of reduction in emergency cardiovascular hospital admissions, we did not observe any evidence of a reduction in admissions with these conditions. We argue that evidence on admission rates should be presented over time as well as statistically. We cannot exclude a modest effect of COVID-19 on admission rates for the serious and treatable emergency conditions studied here.

**Figure 1.**
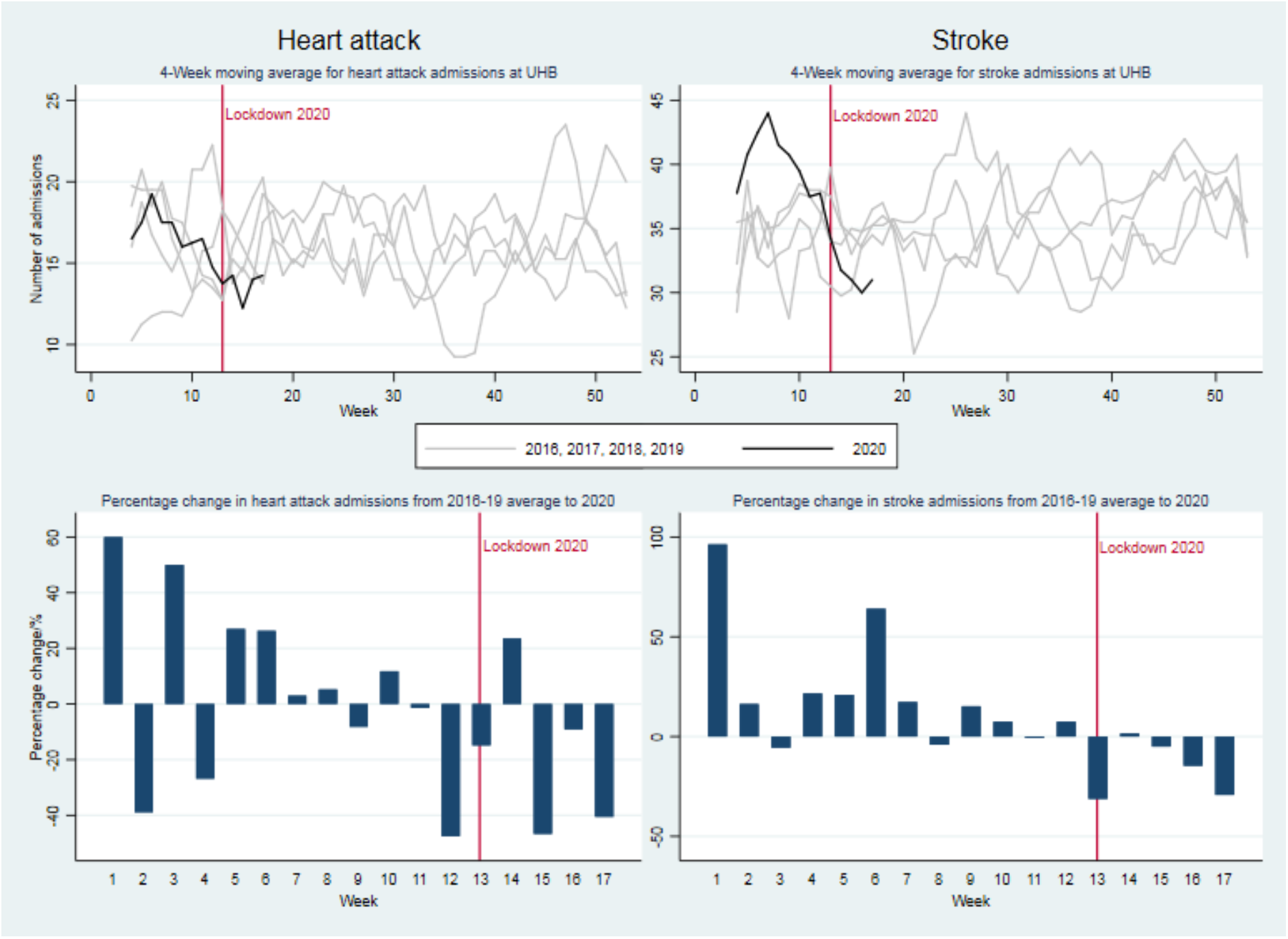
*Top panel: 4-week moving average of weekly admissions for heart attack (left) and Stroke (right). Bottom panel: Percentage change from 2016-2019 average number of admissions to 2020 number of admissions for heart attack (left) and stroke (right). The start of the lockdown period in the United Kingdom is shown by a red line at week 13 (23/04/20)*.

## Data Availability

This work uses data provided by patients and collected by the NHS as part of their care and support at University Hospitals Birmingham NHS Foundation Trust.

## Acknowledgements

This work uses data provided by patients and collected by the NHS as part of their care and support at University Hospitals Birmingham NHS Foundation Trust. It has been approved by University Hospitals Birmingham NHS Foundation Trust, Clinical Audit Registration & Management System and the COVID-19 research facilitation group under application reference [16049].

